# Identification of Genetic Predisposition to Sjögren’s Syndrome by Whole Exome Sequencing

**DOI:** 10.1101/2024.03.14.24304128

**Authors:** Qiwei Guo, Qiaowei Li, Huiqin Lu, Yingying Shi, Jintao Guo, Hao Wang, Qiuxiao Deng, Yihang Li, Yuan Liu, Guixiu Shi, Shiju Chen

## Abstract

A comprehensive understanding of the genetic predisposition associated with the initiation and development of Sjögren’s syndrome (SjS) is imperative. This would not only enrich our knowledge of the pathogenesis underlying this autoimmune disease but also address the long-standing clinical challenges of more timely diagnosis and effective treatment to retain organ function and improve prognosis. In this study, we used whole exome sequencing analysis of 50 patients with SjS to investigate the predisposing variants, genes, and their associated biological functions. Hundreds of predisposing genes were identified, and numerous biological processes and pathways were highlighted; suggesting a heterogeneity of genetic predisposition to SjS. Female patients carrying a greater number of enriched variants tended to have higher levels of serum IgG and corresponding systemic involvement, demonstrating the pivotal role of genetic predisposition in the pathogenesis of SjS. Biological function analysis indicated that a subset of SjS and neuropathies may share a similar genetic predisposition. Our results showed that extracellular matrix-receptor interactions, macrophage-associated biological functions, and motor proteins may play important roles in the pathogenesis of SjS, and macrophage-associated biological functions may be associated with early onset SjS in female patients. Furthermore, the identification of highly enriched variants in the patient cohort provides the possibility of advancing the diagnosis of SjS. In conclusion, our study provides an extensive framework for analysis of the genetic predisposition to SjS which can facilitate further focused and in-depth investigation of the pathogenetic mechanisms of specific genes, biological processes, and pathways; thereby contributing to the pathophysiology, diagnosis, and therapeutics of SjS.

## 1. Introduction

Sjögren’s syndrome (SjS) is a systemic autoimmune disease that affects approximately 0.1–0.5% of the general population, and more than 90% of patients are women [1]. It is characterized by the disorganization and chronic inflammation of the exocrine glands, particularly the salivary and lacrimal glands, resulting in glandular dysfunction and sicca symptoms [2]. Heterogeneous extraglandular manifestations with multi-organ involvement, such as fatigue, synovitis, arthralgia, neuropathy, vasculitis, and interstitial lung disease, are also common in SjS [2]. Furthermore, approximately 5% of patients with SjS also develop non-Hodgkin lymphoma [3].

The pathogenesis of SjS is heterogeneous and remains to be fully elucidated. Generally, SjS initiates and develops when individuals with a genetic predisposition are exposed to environmental or hormonal factors [4]. In affected glands, both innate and adaptive immunity can be activated, causing inflammation and infiltration of immune cells; primarily T and B cells [5]. The pathogenic interplay between epithelial and immune cells can result in glandular tissue destruction, B-cell hyperactivity, formation of ectopic germinal centers, and local production of autoantibodies [6]. A comprehensive understanding of the genetic predisposition involved in the initiation and development of SjS is imperative as it not only enriches our knowledge of the pathogenesis underlying this disease but also addresses the long-standing clinical challenges of more timely diagnosis and effective treatment to retain organ function and improve prognosis [4].

In the past decade, several studies have generated initial insights into the genetic predisposition to SjS. Various predisposing human leukocyte antigen and non-human leukocyte antigen loci in different population backgrounds have been identified by genetic association analysis, a methodology frequently used for common diseases studies that focuses on common variants with allele frequencies ≥ 0.01 [7–11]. However, with the development of high-throughput sequencing, rare and deleterious variants with allele frequencies < 0.01 have been demonstrated to be more highly penetrant contributors to common diseases and have a strong impact on the predisposition to disease in individual patients [12, 13]. Johar et al. identified several rare functional variants in multiple autoimmune syndromes and SjS using whole exome sequencing (WES); however, the sample size was limited (eight patients with SjS patients and other autoimmune diseases and four patients with SjS patients but no other autoimmune diseases), and only homozygous and compound heterozygous variants were analyzed [14]. Subsequently, Wang et al. used WES to analyze rare immune-related variants in 31 families (nine SjS, eleven systemic lupus erythematosus, six rheumatoid arthritis, and five mixed autoimmunity) and identified that the T cell receptor signaling pathway constitutes a shared genetic predisposition for these autoimmune diseases. However, non-immune-related genes were not analyzed [15]. Qi et al. demonstrated that the adenosine monophosphate-activated protein kinase signaling pathway and cell adhesion molecules are involved in the genetic predisposition to SjS, based on WES analysis of 12 patients with SjS (seven individuals from three pedigrees and five sporadic cases) [16]. However, our understanding of rare variant-based genetic predisposition to SjS is limited, and studies containing a larger cohort of sporadic cases are needed to delineate a more comprehensive profile.

In this study, we conducted WES analysis of 50 patients with SjS, most of whom were sporadic SjS cases, to investigate the predisposing variants, genes, and their associated biological functions; aiming to expand our knowledge of the pathogenesis of SjS and facilitate timely diagnosis and effective treatment of this autoimmune disease.

## 2. Materials and Methods

### 2.1. Patients and samples

Fifty patients who fulfilled the American College of Rheumatology/European League Against Rheumatism classification criteria for SjS were enrolled in the Department of Rheumatology and Clinical Immunology, First Affiliated Hospital of Xiamen University, China [6]. According to the available data, 40 patients were diagnosed with primary SjS, whereas 10 patients were diagnosed with associated SjS (concurrence of SjS with other autoimmune diseases). Currently, there is a lack of sufficient evidence to distinguish primary from associated SjS precisely, either from a clinical or biological perspective [1]; therefore, we analyzed these groups of patients non-distinctively. Except for two pairs of first-degree relatives, the patients were consanguineously unrelated. One milliliter of ethylenediaminetetraacetic acid-anticoagulated peripheral blood was collected from each patient for genomic DNA extraction. The patient information is listed in Supplemental Table 1.

### 2.2. Whole exome sequencing and variant output

Genomic DNA was extracted from 400 μL of ethylenediaminetetraacetic acid-anticoagulated peripheral blood using the Super/HF16 Plus DNA Extraction System (MagCore, Xiamen, China) according to the manufacturer’s instructions. Subsequently, the DNA samples were analyzed using a commercial WES service (Novogene, Beijing, China). Briefly, the exome sequences were enriched from 400 ng of genomic DNA using a liquid capture system (Agilent SureSelect Human All Exon V6; Agilent Technologies, Santa Clara, CA, USA), according to the manufacturer’s protocol, and sequenced on the NovaSeq 6000 platform (Illumina, San Diego, CA, USA) to generate 150 bp paired-end reads. The mean exome coverage was above 100×, allowing examination of the selected region with sufficient depth to accurately match more than 99.5% of the targeted exome.

After quality control, high-quality reads were mapped to the human reference genome (hg38) using the Burrows Wheeler Aligner [17]. Subsequently, SAMtools was used for calling single nucleotide variants (SNVs) and insertions and deletions (indels) based on the following filter parameters: mapping quality ≥ 30, quality by depth ≥ 4, and Phred-scaled quality score of variant ≥ 20 [18]. Called SNVs and indels were annotated with ANNOVAR to itemize bioinformatic content, such as the genomic location of the variant, amino acid changes, minor allele frequency (MAF), and *in silico* pathogenicity prediction [19].

### 2.3. Pipeline of variant filtering

Owing to the unsatisfactory accuracy of current WES for indel detection [20], only SNVs were analyzed in this study to reduce the impact of the innate error of WES and increase the specificity of candidate variant filtering. Genes in highly homologous genomic regions that pose technical hurdles for variant analysis were also excluded [21].

A stepwise filtering pipeline was used to identify rare pathogenic variants (Fig. 1). Briefly, variants causing missense, frameshift, or nonsense mutations or splicing changes were retained, and rare variants were further retrieved according to MAF data in three multi-population databases (gnomAD, dbSNP, and 1000 Genomes) and three East Asian population databases (ChinaMAP, jMorp, and KRGDB) [22–26]. Finally, rare variants were filtered by *in silico* pathogenicity prediction using four algorithms, SIFT, PolyPhen-2, Mutation Taster, and CADD, to prioritize the possible pathogenic variants.

**Figure 1.**
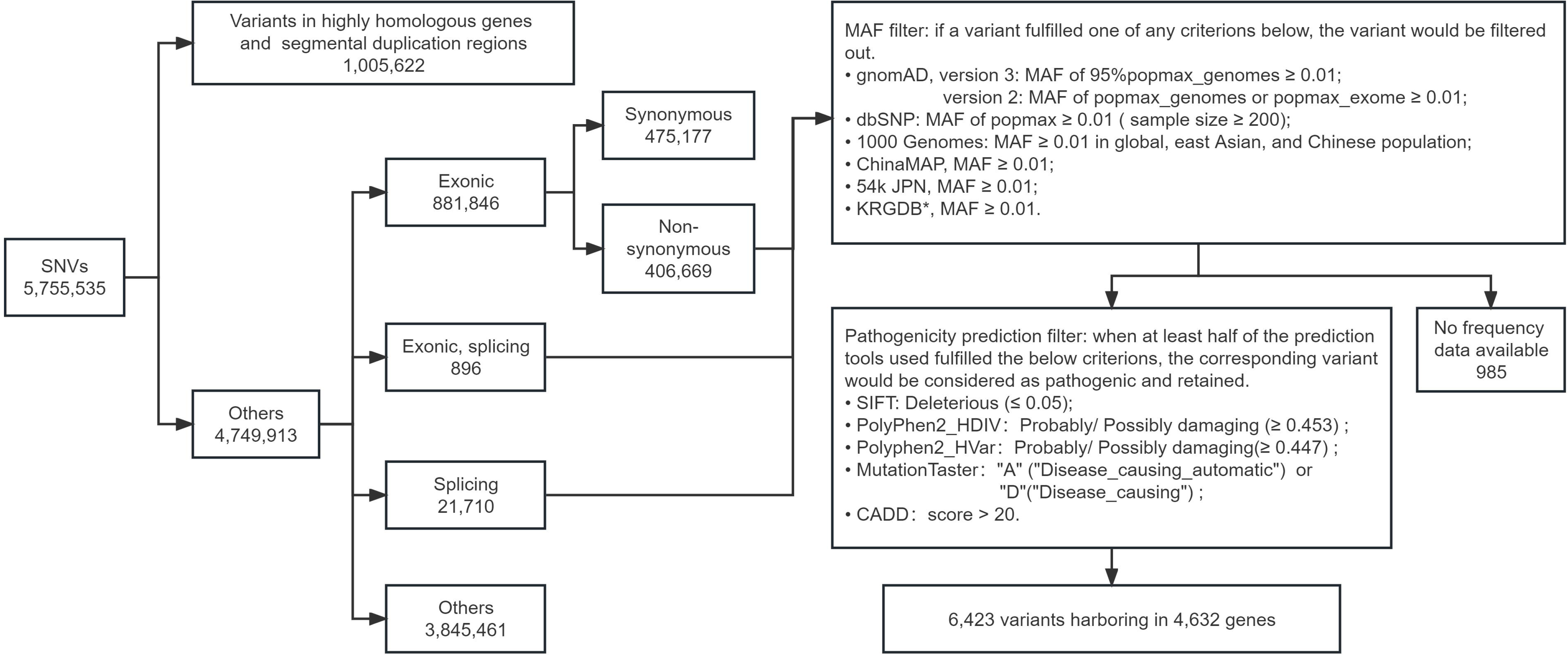
Filtering pipeline for pathogenic rare variants. *Due to the provisional close of KRGDB, the variants with only one allele count were not examined for their minor allele frequencies in this database.

### 2.4. Variant validation by Sanger sequencing

Sanger sequencing is considered the gold standard for validation of candidate variants identified by WES. Therefore, DNA samples with target variants, and two control samples without target variants, were amplified with the corresponding sequencing primers (Supplemental Table 2) on a T100 Thermal Cycler (Bio-Rad, Hercules, CA, USA). The PCR conditions are described in detail in Supplementary Table 3. After amplification, the PCR products were sequenced, using Sanger sequencing, by a commercial sequencing service (Sangon, Shanghai, China).

### 2.5. Rarity validation by real-time PCR-based genotyping

Once the target variants were validated by Sanger sequencing, their rarity in the general population was confirmed using real-time PCR-based genotyping. Briefly, 100–300 genomic DNA samples from random female volunteers, equivalent to 200–600 alleles, were examined using real-time PCR in which hydrolysis probes were designed to identify target variants. Sanger sequencing-validated patient samples were used as positive and negative controls. The primers and hydrolysis probes are listed in Supplemental Tables 2 and 4, respectively.

### 2.6. Functional enrichment and pathway analysis

To determine the enriched biological processes and pathways associated with the candidate genes, The Database for Annotation, Visualization and Integrated Discovery (http://david.abcc.ncifcrf.gov) was used to perform Gene Ontology (GO) enrichment and Kyoto Encyclopedia of Genes and Genomes (KEGG) pathway analyses [27]. GraphPad Prism version 9 (GraphPad, Boston, MA, USA) was used to visualize the enriched GO terms and KEGG pathways.

### 2.7. Statistical analyses

Statistical analysis was performed using SPSS statistical software version 25.0 (IBM, Armonk, NY, USA). Pearson’s (parametric) or Spearman’s (non-parametric) correlation was performed to determine the correlations between the number of carried variants and different clinical indices.

### 2.8. Ethics statement

Signed informed consent was obtained from each participant, and all participants agreed to the use of their data for research purposes. This study was approved by the Research Ethics Committee of the First Affiliated Hospital of Xiamen University, China.

## 3. Results

### 3.1. Overview of candidate variants and genes

After filtering, 6,424 variants in 4,632 genes were identified. The allele count of a specific variant increased as the number of variants decreased (Fig. 2A). To confirm the accuracy of WES and our variant filtering pipeline, the 11 most enriched variants with allele counts of four and five were analyzed further using Sanger sequencing and real-time PCR-based genotyping. As shown in Supplemental Fig. 1, Sanger sequencing confirmed all 11 variants identified by WES. Examination of our sample cohort from the general population revealed that the allele frequencies of most variants (10 of 11) were lower than 0.01. However, the allele frequency of a *TRADD* variant was slightly higher than 0.01 (Fig. 2B), therefore, this was excluded from subsequent analyses. Our results suggest that the majority of candidate variants are rare in the general population, thus validating the variant filtering pipeline.

**Figure 2.**
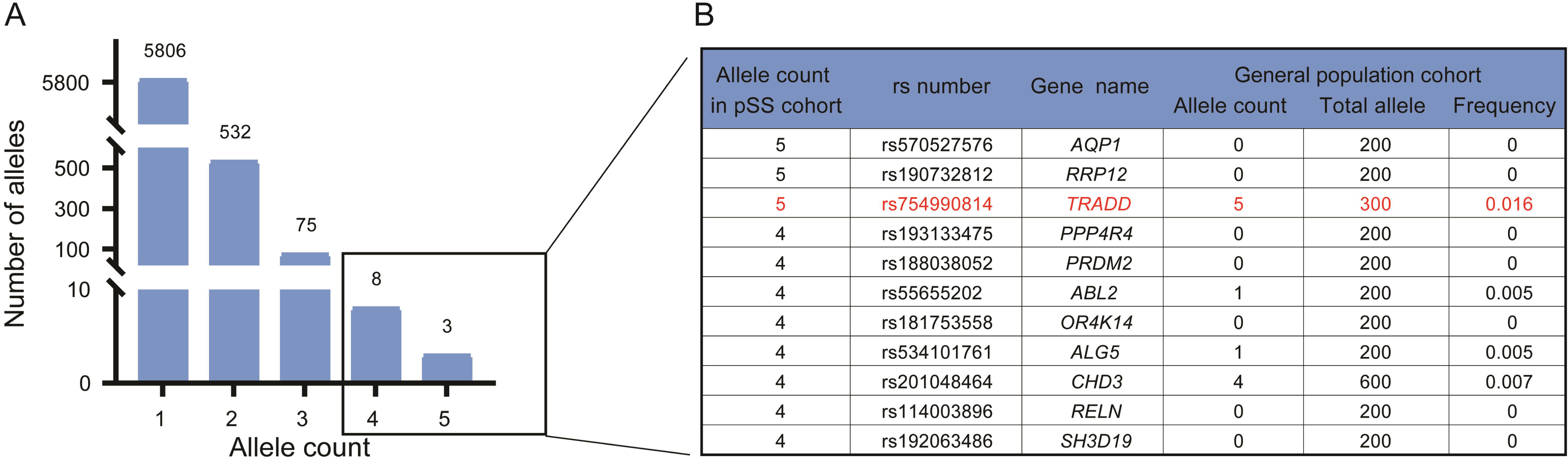
Validation of the variant filtering pipeline. (A) Numbers of alleles with different allele counts. (B) The candidate variants were rare in healthy volunteer cohort.

Most variants were heterozygous. When two or more heterozygous variants are detected in a single gene of a specific patient, both alleles of the gene can be affected in a compound heterozygous manner. Our results showed that 80 genes in 41 patients could be affected by compound heterozygous variants (Supplemental Table 5). In addition to the heterozygous variants, eight variants were homozygous and located in eight genes from six patients (Supplemental Table 6).

Among the 4,632 candidate genes, 24 were identified as predisposing genes for SjS in previous studies, among which *RELN* was most frequently affected (Supplemental Fig. 2).

### 3.2. Functional analysis of enriched genes

Considering that the presence of many variants with only one allele count could be a random rather than an enriched event in patients with SjS, this subset was excluded from further analysis to increase analytical specificity. Therefore, 617 variants with allele counts ≥ 2 (enriched variants) and 601 genes (enriched genes) were subsequently analyzed for their biological functions (Supplemental Tables 7).

Our results showed enrichment of several biological processes, including nervous system-associated biological processes, including nervous system development, single-organism behavior, cell projection organization, and neurological system process; cilium, including the motile and non-motile cilium, organization; immunological processes, including macrophage cytokine production, receptor-mediated endocytosis, and regulation of I-kappaB kinase/NF-kappaB signaling; process of female sex differentiation; and photoreceptor cell differentiation (Fig. 3A and Supplemental Table 8).

**Figure 3.**
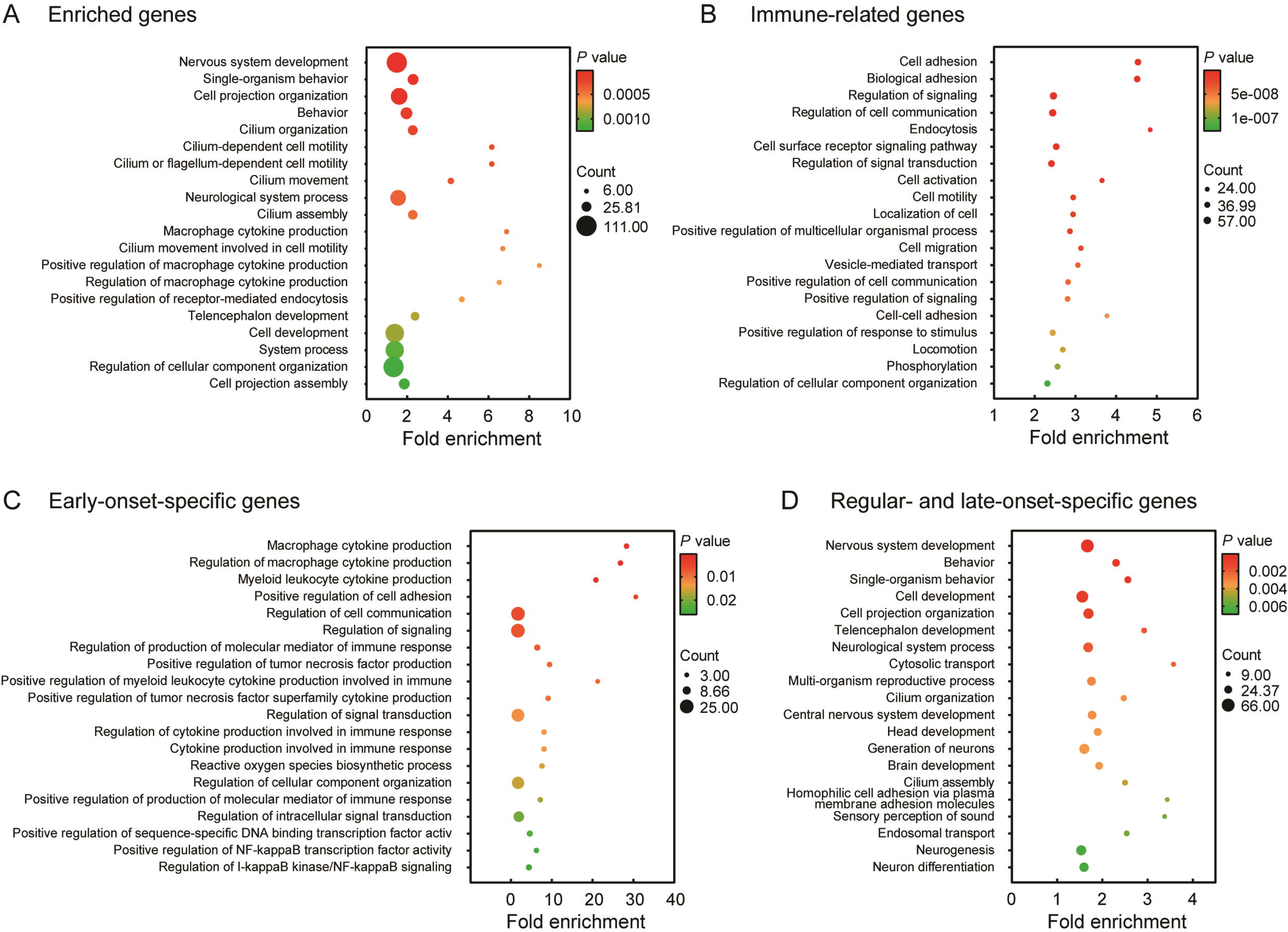
Top 20 enriched biological processes for different gene sets. (A) Enriched biological processes for the genes enriched in patients with SjS, (B) immune-related genes among the enriched genes, (C) early-onset-specific genes, and (D) regular- and late-onset-specific genes.

Several pathways were enriched for the 601 enriched genes, most of which were immune-related (Fig. 4A and Supplemental Table 9); underlining the contribution of an immune-related genetic predisposition to SjS. Among these enriched pathways, the extracellular matrix (ECM)-receptor interaction was the most enriched (*P*=2.24E-06).

**Figure 4.**
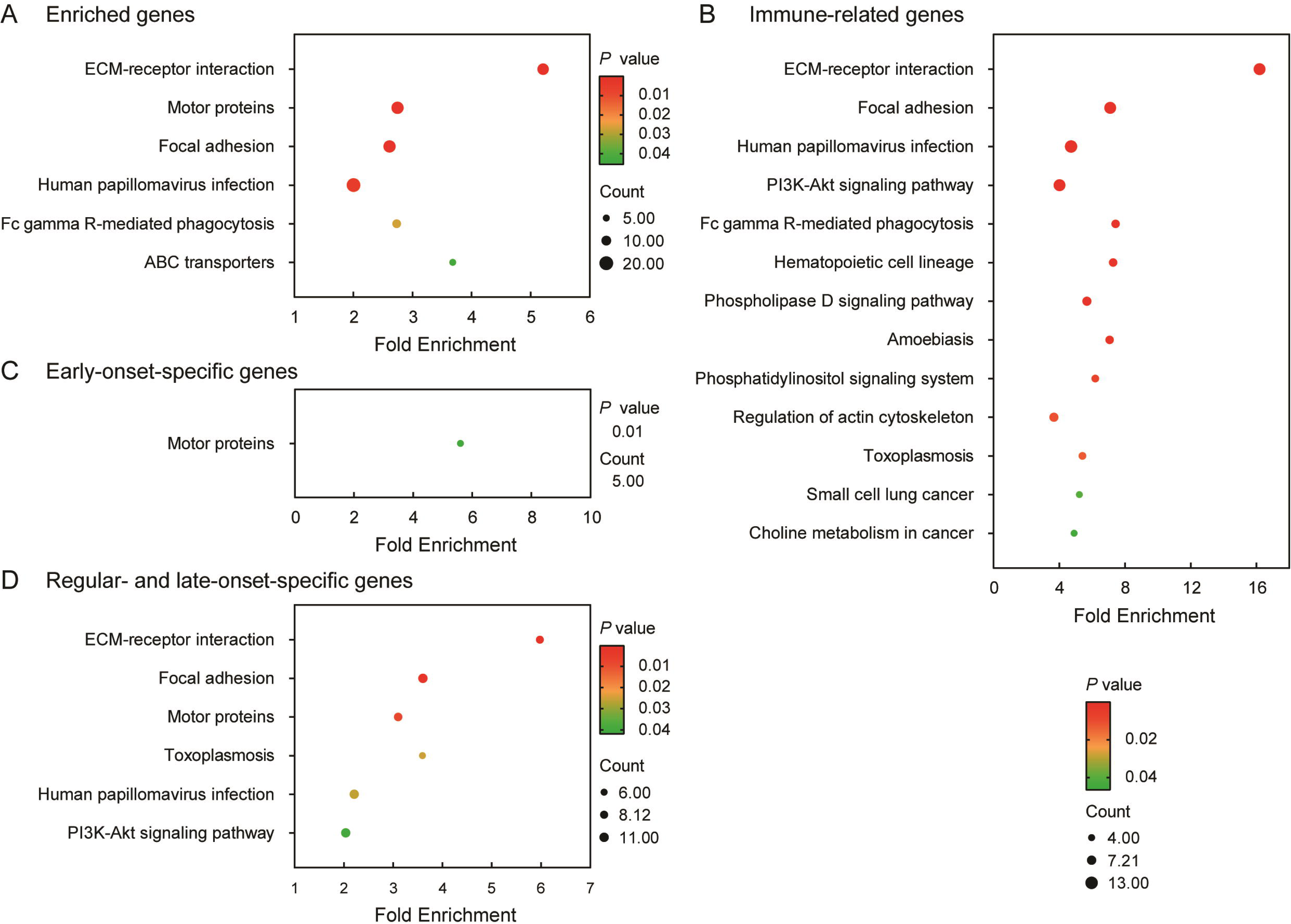
Significantly enriched KEGG pathways (*P*<0.05) for different gene sets. (A) Enriched pathways for the genes enriched in patients with SjS, (B) immune-related genes among enriched genes, (C) early-onset-specific genes, and (D) regular- and late-onset-specific genes.

### 3.3. Functional analysis of immune-related genes

According to the immune-related genes list in the innateDB [28], 121 genes were immune-related among the 601 enriched genes (Supplemental Table 10). To further evaluate the genetic predisposition of immune factors in SjS, we performed functional analysis of the immune-related gene set.

Notably, cell adhesion was strongly enriched (*P*=1.95E-18); and the regulation of signaling and associated cellular activities, such as cell activation, endocytosis, and cell migration, were also enriched. Furthermore, nervous system development was associated with this subset of genes. (Fig. 3B and Supplementary Table 11).

KEGG pathway analysis (Fig. 4B and Supplemental Table 12) showed that the ECM-receptor interaction pathway was strongly enriched (*P*=1.13E-10), which is consistent with the results from the full 601 enriched genes (Fig. 4A). The Fc gamma receptor (FcγR)-mediated phagocytosis and associated signaling pathways were also enriched.

### 3.4. Functional analysis of genes affected by homozygous variants and potentially compound heterozygous variants

Compared to heterozygous variants, homozygous and compound heterozygous variants could result in impaired function of both alleles, and the affected genes would have a closer relationship with the pathogenesis of SjS. As shown in Supplemental Table 13, the main enriched biological processes were similar to those of the enriched genes (Supplemental Table 9), including regulation of homeostatic processes, especially calcium homeostasis; system processes, including neurological and muscle system processes; cell adhesion; and nervous system development, including glial and photoreceptor cell development.

KEGG pathway analysis revealed enrichment in the calcium signaling and pyrimidine metabolism pathways (Supplemental Table 14).

### 3.5. The presence of a greater number of enriched variants in female patients could promote autoimmunity and thus increase the risk of systemic involvement

In our patient cohort, 23 female patients did not receive any treatment at the time of enrolment (Supplemental Table 1), enabling us to investigate the effects of genetic predisposition on clinical indices. Statistical analyses were performed to evaluate the association between the number of enriched variants in an individual and various clinical indices. As shown in Table 2 and Fig. 5A and B, female patients who carried a greater number of enriched variants tended to have higher levels of IgG in their serum as well as higher European League Against Rheumatism Sjögren’s syndrome disease activity index (ESSDAI) scores. Furthermore, higher serum IgG levels were associated with higher ESSDAI scores (Fig. 5C). These results suggest that a higher number of predisposing variants in female patients could promote autoimmunity, thereby increasing the risk of systemic involvement.

**Figure 5.**
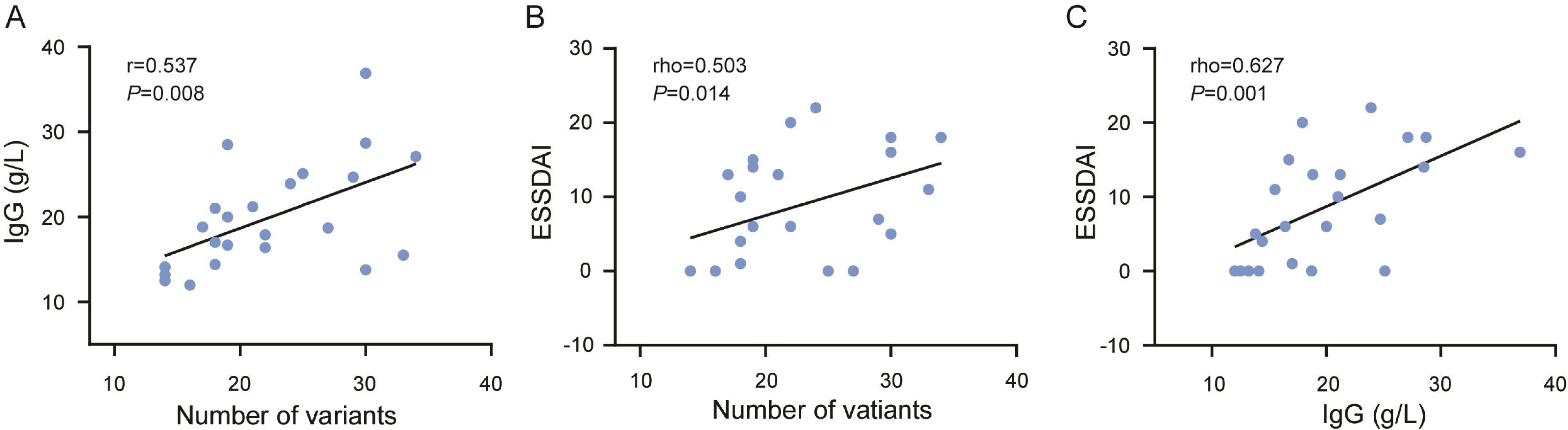
Carrying a greater number of enriched variants could promote autoimmunity and thus increase the risk of systemic involvement in female patients. (A) Associations between the number of variants in an individual and serum IgG levels. (B) Associations between the number of variants in an individual and ESSDAI scores. (C) Associations between serum IgG levels and ESSDAI scores. IgG, immunoglobin G; ESSDAI, European League Against Rheumatism Sjögren’s syndrome disease activity index.

### 3.6. Macrophage-associated processes are associated with early onset of disease in female patients

Patients with an onset age of 35 or younger were defined as having early-onset SjS, whereas the others were defined as having regular- or late-onset SjS; according to the criteria from previous studies [29, 30]. In this context, 10 and 36 female patients were considered early onset and regular- and late-onset patients, respectively (Supplemental Table. 1). Eighty-one enriched genes were found to be specific to the early-onset subset, whereas 318 enriched genes were specific to the regular- and late-onset subset (Supplemental Fig. 3 and Supplemental Tables 15 and 16). Functional analysis showed that immune-associated processes, particularly macrophage-associated processes, were enriched in the early onset-specific gene set; whereas biological processes of nervous system development were enriched in the regular- and late-onset-specific gene sets (Fig. 3C and D, Supplemental Tables 17 and 18). Similarly, different enriched pathways were identified for the different age of onset subsets (Fig. 4C and D; Supplemental Tables 19 and 20).

## 4. Discussion

In this study, we investigated the genetic predisposition to SjS by evaluating enriched rare variants derived from WES data of 50 patients with SjS. By examining the allele frequencies of the 11 most enriched variants in our general population cohort (Fig. 2), we validated our filtering pipeline’s ability to identify rare variants; which was a prerequisite for achieving the goal of this study. The enrichment of hundreds of rare variants in the SjS cohort, and the discovery that female patients who carry a greater number of enriched variants tend to manifest higher levels of serum IgG and corresponding disease activity, demonstrate the pivotal role of genetic predisposition in the initiation and development of this autoimmune disease.

Based on the functional analysis of enriched variant-associated gene sets, we identified numerous associated biological processes and pathways (Fig. 3 and 4, Supplemental Tables 8–20); suggesting the heterogeneity of the genetic predisposition in SjS and providing an extensive framework for further focused and in-depth analyses to advance our knowledge of the pathogenesis of SjS.

A subset of SjS-associated predisposing genes was also involved in nervous system-associated biological processes, such as nervous system development and neurological system processes; suggesting an overlap in the genetic predisposition to SjS and neuropathies. Notably, this result was also identified in genes affected by homozygous and potentially compound heterozygous variants, which are considered closely related to the pathogenesis of SjS. An increasing number of studies have shown that neurological involvement in SjS has been previously underestimated or under-diagnosed [31–33]. More recently, Seeliger et al. demonstrated that nearly half of patients with SjS suffered from neurological involvement, including muscular, central, and peripheral nervous system, and this subset of patients have a different clinical phenotype from that of patients without neurological involvement [31]. These findings support our hypothesis that a subset of SjS and neuropathies share a similar genetic predisposition and suggest that a specific phenotype could be shaped by this underlying genetic predisposition. Seeliger et al. demonstrated that neurological involvement in SjS was associated with an older age of onset. This is consistent with our finding that genes involved in nervous system development were enriched in female patients with regular- and late-onset rather than early onset SjS (Fig. 3D and Supplemental Table 18), further suggesting the importance of genetic predisposition in phenotype shaping. Previously, neuropathies in SjS had been suggested to be secondary to the vasculopathy or glandular autoimmunity [34, 35]. However, along with the high incidence of neuropathies in SjS, the positive relationship between neuropathies and autoimmunity was challenged [33]. Furthermore, the therapeutic effects of immunosuppressants on SjS-associated neuropathies are limited in some cases [35]. These findings suggest that neuropathies, at least in part, result from a genetic predisposition rather than secondary injury to glandular autoimmunity [31, 33]. One of the putative etiologies underlying the hypothesis of shared genetic predisposition is that specific predisposing genes could have multiple functions that simultaneously regulate the homeostasis of the nervous system, immune system, and salivary/lacrimal glands. Several lines of evidence associated with our identified enriched genes support this hypothesis. For example, the products of the four most enriched candidate genes, AQP1, ABL2, CHD3, and RELN, participate in inflammatory activation, cell adhesion and migration, wound healing, and organ regeneration [36–41]. In addition, they play important roles in the nervous system and are associated with central and periphery neuropathies [39, 42–46]. Motor proteins, which are the products of another subset of enriched genes, are essential for axonal transportation and function at immunological synapses [47, 48].

Macrophage-associated enriched variants can simultaneously affect the homeostasis of the nervous system and salivary/lacrimal glands by altering the function of tissue-resident macrophages [49, 50]. Another putative etiology underlying the hypothesis of shared genetic predisposition is that because the immune responses and gland morphogenesis, vascularization, and functionality can be modulated by the nervous system, its dysregulation disrupts homeostasis of the immune system or salivary/lacrimal glands [51–56].

Variants in genes encoding various ECM proteins, such as laminin, collagen, reelin, fibronectin, and thrombospondin, were enriched in our study; therefore, the ECM-receptor interaction was identified as an important player in the genetic predisposition to SjS (Fig. 3 and 4). The ECM provides various biomaterial scaffolds which cells can sense, adhere to, and remodel to control cell shape, motility, and signaling; and is essential for a broad array of biological processes such as development, hematopoiesis, immune response, wound healing, and organ regeneration [57]. In particular, several ECM-associated genes encode basement membrane (BM) proteins, such as LAMA2, LAMB1, LAMC3, FN1, COL6A3, COL9A2 [58], were identified in the present study. The BM is an ECM lining the mesenchymal-epithelial junction, which is physiologically pivotal for branching morphogenesis, cellular polarity of the salivary gland, and migration and differentiation of progenitor cells via interaction with associated receptors; thereby maintaining glandular homeostasis [59, 60]. Disorganization of the BM owing to the pathogenic variants could not only result in the defective structure and function of salivary gland but also be associated with infection and mononuclear cell infiltration in SjS [61, 62] (Fig. 6). In the pathological state, ECM components can serve as damage-associated molecular patterns (DAMPs) that exacerbate and perpetuate inflammatory cascades and may provide a novel source of chronic B cell activation in SjS [63].

**Figure 6.**
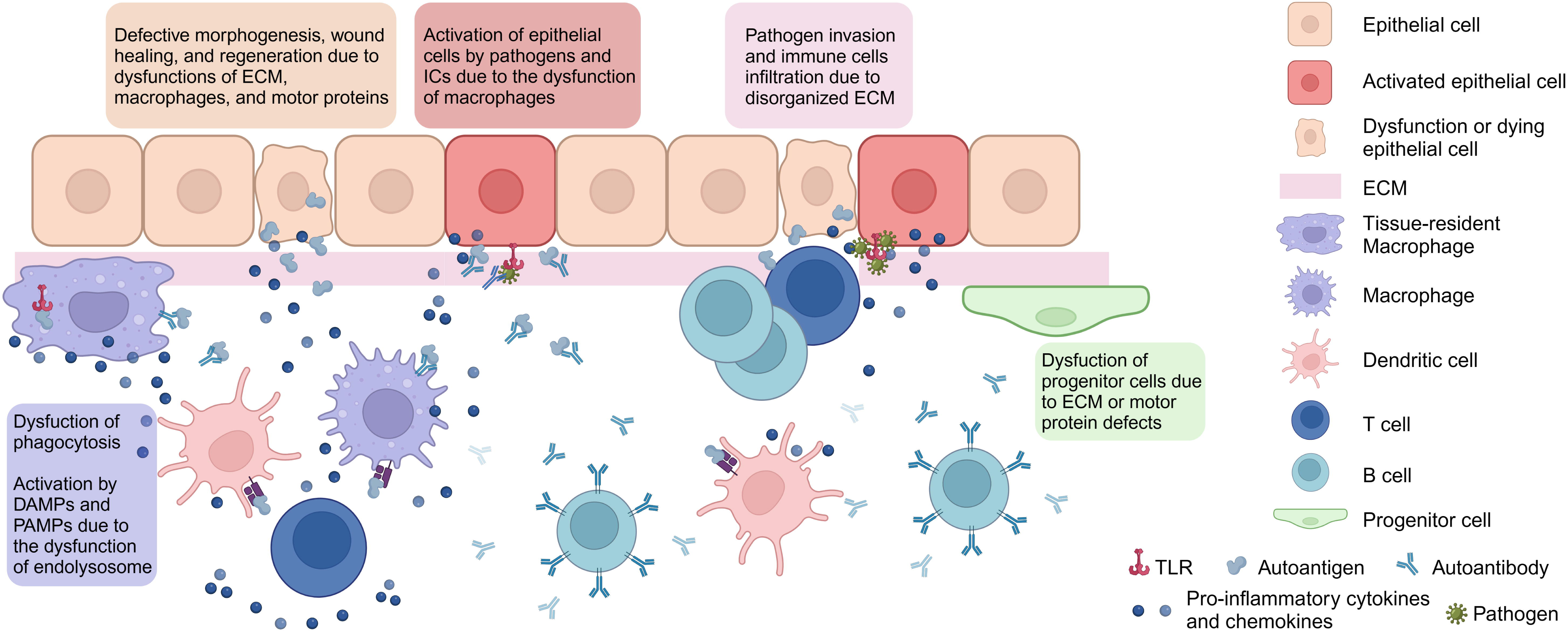
Putative pathogenesis of Sjögren’s Syndrome associated with genetic predisposition. ECM, extracellular matrix; ICs, immune complexes, TLR, toll-like receptor; DAMPs, damage-associated molecular patterns; PAMPs, pathogen-associated molecular patterns.

Macrophages are suggested to be an important immunological player in SjS, particularly in early onset, which manifests higher levels of autoimmunity and is associated with poor prognosis that should be monitored more strictly [29, 64]. Macrophages and their corresponding cytokines have been identified in the inflammatory glands of both patients with SjS and animal models and are positively correlated with the severity of glandular morbidity [50, 65, 66]. They are heterogeneous cell populations that are ubiquitous in all organs and serve as active participants in developmental processes and immune responses to maintain organic integrity and homeostasis [49, 50]. For example, macrophages remove cellular debris, apoptotic cells, senescent cells, invading pathogens, and immune complexes (ICs) by phagocytosis; thereby playing an important role in host defense, morphogenesis, and tissue homeostasis. In our study, FcγR-mediated phagocytosis and related signal pathways were associated with the genetic predisposition to SjS. FcγRs are a group of key receptors involved in the engulfment of opsonized microorganisms and immune complexes [67]. FcγR signaling for phagocytosis involves the activation of serial signal cascades, including PI3K-Akt, phospholipase D, NF-κB, JNK, and MAPK signaling pathway, to reorganize the cytoskeleton and induce an inflammatory response [68]. One putative pathogenic association between dysregulated FcγR-mediated phagocytosis and SjS is that inefficient clearance of invading pathogens in glands could activate epithelial and innate immune cells by other receptors, such as toll-like receptors; thereby inducing tissue injury and generating inflammatory milieu for autoimmunity [69] (Fig. 6). Another putative pathogenic association between dysregulated FcγR-mediated phagocytosis and SjS suggests the inefficient clearance of ICs. Physically, ICs are efficiently eliminated by phagocytes to prevent their accumulation. However, in the pathological state, such as defective phagocytosis resulting from loss-of-function variants, ICs could be deposited on tissues and initiate several immune cascades; thus causing glandular and extra-glandular injuries [70] (Fig. 6). Several studies have indicated the presence of ICs, which can be formed by autoantigens and autoantibodies, in the salivary glands, blood vessels, glomeruli, and serum of patients with SjS [5, 71, 72]. Furthermore, endogenic defects in phagocytosis mediated by other receptors are associated with autoimmune diseases [73]. In addition to dysregulated phagocytosis, impaired phagolysosome function can also result in the accumulation of residual DAMPs or pathogen-associated molecular patterns and trigger local inflammation through pattern recognition receptors. In this study, a PLD3 p.T430I variant was found in a early-onset female patient that drastically impaired its enzymolytic activity for auto-DNA in the phagolysosome of macrophages (unpublished data). This could contribute to the initiation of SjS when triggered by a local overload of auto-DNA since PLD3 has been recently recognized as an important nuclease that limits auto-inflammation by eliminating DNA and RNA in the endolysosome [74]. In addition to their phagocytic ability, macrophages also participate in glandular morphogenesis, wound healing, regeneration, and disinfection by releasing biochemical factors such as trophic factors, coagulation factors, matrix metalloproteases, cytokines, and chemokines [75–79]. Therefore, loss-of-function variants could impair the above-mentioned process, thus interrupting glandular homeostasis. In contrast, macrophages with gain-of-function variants can produce dysregulated inflammatory cytokines, such as interleukin-1 and tumor necrosis factor, to facilitate autoimmunity and cause glandular and peripheral nerve injuries [80, 81]. Notably, several studies have shown that predisposing variants in Alzheimer’s Disease are associated with genes expressed in macrophages [82, 83], which could partly explain why an increased risk of Alzheimer’s Disease is found in patients with SjS and further supports the hypothesis of shared genetic predisposition between SjS and neuropathies [84]. However, the specific contribution of macrophages to early onset SjS requires more comprehensive and in-depth investigation.

Motor proteins, including myosin, dynactin, kinesin, and axonemal dynein, were enriched in the patients with SjS. These proteins function within the cytoskeleton and are essential for various biological processes. Myosin is involved in the cytoskeleton reorganization, which is important for the development of the salivary gland, migration of progenitor and immune cells, cell adhesion, formation of immune synapse, axonal transport, and phagocytosis [48, 85–87]. The dynein/dynactin complex and kinesin transport cytoplasmic cargo, such as mitochondria, viral particles, synaptic vesicles, lysosomes, signaling endosomes, autophagosomes, and microtubules, in a retrograde and anterograde manner, respectively [88, 89]. Disruption of cargo transportation by defects in these motor proteins could result in a range of neurodevelopmental and neurodegenerative diseases, dysfunction of lacrimal acinar cells, disturbed innate and acquired immune response to viral infection, as well as enhancing the activity of NF-κB; a well-known transcriptional factor involved in the pathogenesis of SjS [47, 90–93]. Furthermore, the disruption of dynein and dynactin can impair autophagy and cause defects in the salivary glands, muscles, and nervous system in a motor-independent manner [94]. Overall, the involvement of motor proteins in the genetic predisposition to SjS provides persuasive evidence for the hypothesis of a shared genetic predisposition between SjS and neuropathies. Interestingly, several genes encoding axonemal dyneins were identified in this study (Supplemental Table 7). Axonemal dyneins are foundational proteins of cilia that drive cilium beating, which is essential for various biological processes such as cerebrospinal fluid circulation, mucociliary clearance defense, and spermatozoa motion [95–97]. Therefore, primary ciliary dyskinesia caused by pathogenic variants of the axonemal dynein genes can manifest in various systems [98, 99]. However, this is the first time that axonemal dyneins have been associated with SjS. One putative cause is that ciliary dyskinesia impairs the mucociliary clearance defense, which increases the risk of respiratory infection that is associated with both seropositive and seronegative SjS [96, 98, 100]. Furthermore, axonemal dyneins may also have cilium-independent functions. For example, DNAH5 is required for sound sensation in *Drosophila* larvae and is associated with myeloma [101, 102]. However, the role of axonemal dyneins in the pathogenesis of SjS requires further investigation.

Numerous predisposing genes, biological processes, and pathways were also identified in our study, such as female sex differentiation, cell adhesion, and PI3K-Akt/mTOR signaling, which deserve further investigation for their pathogenic roles in SjS. Genes affected by homozygous and potentially compound heterozygous variants, those frequently identified in different genetic predisposition studies (Supplemental Fig. 2), and their associated biological processes and pathways would be of considerable interest in future studies. For example, *RELN* was frequently affected in our patient cohort and has been identified as a predisposing gene for SjS in previous genome-wide association studies or WES-based analyses [9, 103].

The identification of genetic predisposition, especially highly enriched variants, provides the possibility to advance the diagnosis of SjS. For example, nine variants were identified in 24 of 50 patients with SjS compared to only two of the 100 individuals of the healthy volunteer cohort in our study, suggesting high sensitivity and specificity of these variants for SjS identification (Supplemental Table 21). Larger sample sizes and samples from contrasting diseases, such as systemic lupus erythematosus and rheumatoid arthritis, are required to further evaluate the diagnostic sensitivities and specificities of these candidate variants. However, the diagnostic utility of testing genetic predisposition, whether independently or combined with other phenotype-based measurements [104], would be a promising topic; not only because genetic testing is more stable than serological testing and less invasive than labial salivary gland biopsy but also because it could facilitate the identification of patients at an early stage, which is critical for improving management strategies [105].

The management of SjS is challenging. To date, the vast majority of drugs have failed to demonstrate clinical efficacy based on the outcomes of corresponding randomized placebo-controlled trials; therefore, no drugs have been officially licensed for the treatment of SjS [106]. However, when stratifying patients based on symptoms, the effects of drugs can be positive for specific subgroups of patients [107], and the heterogeneity of SjS pathogenesis is attributable to these findings and highlights the importance of personalized treatment [106]. Our study provides a foundation for developing more personalized therapeutics based on the genetic predisposition that reflects the underlying pathogenesis and for determining the optimal treatment options for subgroups of patients, or even at the individual level for those affected by specific genetic defects. For example, glandular dysfunction and systemic inflammation of patients with *AQP1* variants could be resolved via aquaporin gene therapy [108]; the salivary glands of patients with ECM deficiency could be regenerated by the injection of decellularized ECM hydrogel [109]; and the glandular dysfunction and autoimmunity of patients with *TSC2* variants could be alleviated by seletalisib, a phosphatidylinositol 3-kinase inhibitor [110].

This study was based on an East Asian population, and the genetic predisposition to SjS could differ markedly according to ancestry [9]. Therefore, future studies involving different populations are needed to enable a more comprehensive understanding of the genetic predisposition to SjS. Furthermore, with the development of sequencing technologies and data analysis methodologies, a more accurate analysis of SNVs, indels, copy number variants, and epigenetic changes in whole genomes may further advance our knowledge of genetic predisposition to SjS.

In conclusion, our study provides an extensive framework for understanding the genetic predisposition to SjS based on WES and rare variant analysis and facilitates further focused and in-depth investigation of the pathogenetic mechanisms of specific genes, biological processes, and pathways; thus contributing to the pathophysiology, diagnostics, and therapeutics of SjS.

## Data Availability

All data produced in the present study are available upon reasonable request to the authors

## Acknowledgement

We thank Drs Zhibin Li, Jinxiu Xuan, Yan He, Hongyan Qian, Xinwei Zhang, Yan Li, and Minjie Zhang of the First Affiliated Hospital of Xiamen University for their kind supports to this work.

## Funding

This work was supported by the National Natural Science Foundation of China [grant numbers 82171779] and the Xiamen Municipal Bureau of Science and Technology [grant number 2022XMSLCYX01].

## Declarations of interest

None

---

**Table 1.** Associations between the number of enriched variants in an individual and various clinical indices in female patients.

| Clinical indexes | Case number | <sup>a</sup>r/rho | *P* value |
| --- | --- | --- | --- |
| Onset age | 46 | -0.029 | <sup>b</sup>0.848 |
| Focus score | 22 | 0.176 | <sup>b</sup>0.434 |
| Schirmer's test, left | 21 | 0.029 | <sup>b</sup>0.902 |
| Schirmer's test, right | 21 | 0.244 | <sup>b</sup>0.287 |
| Schirmer's test, mean | 21 | 0.186 | <sup>b</sup>0.419 |
| ANA | 20 | 0.002 | <sup>b</sup>0.992 |
| ACA (IgG) | 23 | 0.141 | <sup>b</sup>0.520 |
| Anti-β2-GP1 Ab (IgG) | 23 | 0.039 | <sup>b</sup>0.860 |
| Anti-SSA Ab | 22 | 0.113 | <sup>b</sup>0.618 |
| Anti-Ro-52 Ab | 22 | 0.268 | <sup>b</sup>0.229 |
| Anti-SSB Ab | 22 | 0.027 | <sup>b</sup>0.905 |
| Anti-TPO Ab | 17 | 0.255 | <sup>b</sup>0.324 |
| IgG | 23 | 0.537 | <sup>c</sup>**0.008** |
| C3 | 23 | -0.032 | <sup>c</sup>0.886 |
| C4 | 23 | -0.149 | <sup>c</sup>0.498 |
| RF | 17 | 0.179 | <sup>b</sup>0.491 |
| ESSDAI | 23 | 0.503 | <sup>b</sup>**0.014** |

<sup>a</sup>r, Pearson's correlation coefficient; rho, Spearman's rank correlation coefficient.
<sup>b</sup>*P* value was calculated with Spearman's rank correlation analysis.
<sup>c</sup>*P* value was calculated with Pearson's correlation analysis.

## Notes

### Competing Interest Statement

The authors have declared no competing interest.

